# Factors Influencing AVF Dysfunction in Patients with Maintenance Hemodialysis: A retrospective study

**DOI:** 10.1101/2021.01.11.20248863

**Authors:** Yu Li, Wenhao Cui, Jukun Wang, Chao Zhang, Tao Luo

## Abstract

**Objective:** To explore factors influencing primary dysfunction of autologous arteriovenous fistula (AVF) in patients with maintenance hemodialysis (MHD).

**Methods:** This is a retrospectively study. One hundred and twenty-one patients who underwent anastomosis for AVF in our hospital Between 1st January 2016 and 31st December 2018 were selected. Seventy-seven patients satisfied the inclusion criteria and were included in the final analyses. Patients were divided into two groups based on the function of vascular access. The comparisons of complete blood count and other blood biochemical parameters were made between two groups. Factors influencing AVF dysfunction were analyzed by multivariate Cox proportional hazard regression model for patients with End stage renal disease (ESRD).

**Results:** There were significant differences in total cholesterol (TC), low density lipoprotein (LDL) and serum phosphorus levels between patency and dysfunction group (P<0.05) of AVFs. Further multivariate COX proportional risk regression showed that hypercholesterolemia and hyperphosphatemia were independent risk factors for AVF dysfunction.

**Conclusions:** Hypercholesterolemia and hyperphosphatemia are independent risk factors for primary AVF dysfunction in patients with MHD.

## 1. Introduction

Advances in medical technology have improved diagnosis and treatment of several diseases and their complications. Nonetheless, mortality due to multiple chronic complications have by far bettered those of other diseases, with chronic kidney disease (CKD) being the most prevalent. Currently, even though kidney transplantation is the most effective treatment for CKD, its application is limited by few available donors. Therefore, maintenance hemodialysis (MHD) remains the best alternative for kidney transplant. Unfortunately, MHD patients require essential lifetime vascular access during dialysis. Autologous arteriovenous fistula (AVF) is the optimal vascular access for MHD. However, possible AVF dysfunction after surgery, particular for prolonged use, has limited its use. The patient had to undergo surgery again if the AVF could not meet the require. This will not only increase the costs, but also lead to the waste of vascular resources. Therefore, the purpose of this paper is to study the relevant factors affecting AVF dysfunction.

We present the following article in accordance with the STROBE reporting checklist.

## 2. Methods

### 2.1 Research object

#### 2.1.1 Selection of study participants

Between 1st January 2016 and 31st December 2018, 121 patients who underwent vascular anastomosis for MHD at the Department of general surgery, Xuanwu Hospital, Capital Medical University were selected for this study.

#### 2.1.2 Inclusion criteria

1. Must have been 18 years or older at the time of fistula establishment.
2. Meant to undergo AVF for the first time.
3. The newly established AVF must be functioning normally, and the blood flow in AVF equal or above 250ml / min.
4. Patients must undergo HMD in our hospital after AVF establishment.
5. All patients were operated by the same doctor.

#### 2.1.3 Exclusion criteria

1. Patients unable to receive telephone follow-up for various reasons.
2. Patients with a short life expectancy due to various causes (e.g., tumors).
3. Patients who underwent renal transplantation, were lost to follow-up, or died.
4. Patients with other causes of AVF dysfunction.

#### 2.1.4 Grouping of patients

A total of 121 patients were included in this study. After assessment, 77 patients fulfilled the inclusion criteria and were included in the study. Patients were divided into patency (n= 47) and dysfunction (n = 30) group based on whether their AVF was patency or not. The protocol of this retrospective designed study conformed to the ethical guidelines of the Declaration of Helsinki (as revised in 2013) and was approved by the Ethics Committee of Xuanwu Hospital (No. L YS [2020]018) with waiver of informed consent.

### 2.2. Research methods

#### 2.2.1 Surgical procedure

First, the necessity and safety of operation were carefully evaluated. Thereafter, the diameter of radial artery and cephalic vein for all patients were examined by ultrasound before operation. End to side forearm cephalic vein-radial artery anastomosis was performed in patients without contraindications under a local anesthesia. Ultrasounds for radial artery and cephalic vein were repeated after surgery, but before dialysis. Regular dialysis was started on non-stenosis patients. The operation was performed by the same doctor.

#### 2.2.1 Assessment for successful operation and dysfunction

AVF was deemed successfully if vascular pulsation was palpable, there were apparent tremor, and the blood flow was above the minimum rate required for dialysis (250ml/min). A functional AVF must be absence of thrombosis, stenosis, hematoma and other serious complications. Briefly, AVF was deemed to have dysfunctioned if it could not supply more than 200 ml/min of blood flow stably, accompanied by weak or occasional tremor or heart murmur and if vascular ultrasound revealed stenosis or thrombosis.

#### 2.2.2 Blood Biochemical tests

The patients underwent regularly blood tests on an empty stomach before dialysis. All laboratory indicators were measured at least 3 months after dialysis initiation, and we collected the most recent data before AVF dysfunction (or before the study if patency). In addition, samples were collected by qualified health professionals.

#### 2.2.3 Observation variables

During the assessment period, we recorded the gender, age at AVF establishment, BMI, primary disease of End stage renal disease (ESRD), preoperative blood pressure, glycosylated hemoglobin (HbA1c), diameter of blood vessels, hemoglobin (HB), albumin (ALB), total cholesterol (TC), total triglycerides (TT), low density lipoprotein (LDL), creatinine (SCr), and preoperative electrolyte condition (procalcitonin PTH, blood calcium, blood phosphorus, calcium phosphorus product). At the same time, AVF patency was also recorded (A blood flow of 250 ml/min or more and adequate for MHD).

#### 2.2.4 Statistical analyses

Continuous data were expressed as mean ± standard deviation. Comparisons between groups were performed by independent t-tests if normal distribution is conformed. Mann Whitney U test was used if normal distribution was not conformed. Categorical variables were reported using frequency counts and percentages, with statistical differences between groups performed using chi-square test. First, identify the variables with difference between patency and dysfunction group by univariate analysis. Second, A cox proportional hazard regression was performed, in which variables with previous differences were include. “Enter” method was adopted to screen variables. It was considered statistically significant if P<0.05. Statistical analyses were performed with SPSS 19.

## 3. Result

### 3.1. Follow-up results and overall patency of fistula

Generally, 121 patients with ESRD were included in our center. Of the 121 patients, 23 were lost to follow-up, 10 received kidney transplant, and 11 died. 77 patients who met the inclusion criteria were included in the analysis. Among them, 53 (68.8%) were male whereas 24 (31.2%) were female. The participants ranged from 29-82 years old, with an average age of (56.5±11.3) years. Diabetic and non-diabetic nephropathy accounted for 47 (61.0%) and 30 (39.0%) of ESRD cases. All AVFs were matured, and the follow-up ranged from 5 to 39 month with a mean of (20.9 ± 10.1) month. Meanwhile, the follow-up period of patients in patency group and dysfunction group was (19.7±9.9) month and (22.8±10.2) months, respectively. Since the follow-up period was non normal distribution, Mann Whitney U test was performed, and there was no statistically significant difference between the patency and dysfunction groups (P = 0.191). During the observation period, 30 (39.0%) patients developed AVF dysfunction whereas 47 (61.0%) patients did not.

### 3.2. Comparison of baseline between patency and dysfunction group

The clinical and demographic characteristics of the entire study cohort are summarized in Table 1. A total of 21 study factors were included and compared between the patency and dysfunction group. P-values of five variables were less than 0.1, which were Gender, Phosphorus, Calcium Phosphorus product, TC and LDL. However, there was no significant difference for other factors. (Table 1).

**Table 1.** Baseline of Patency and Dysfunction Group.

|  | Overall<br>(N=77) | Patency<br>(N=47) | Dysfunction<br>(N=30) | Sig |
| --- | --- | --- | --- | --- |
| Follow up time (Month) | 20.9 ± 10.1 | 19.7 ± 9.9 | 22.8 ± 10.2 | 0.191 |
| Female (%) | 24 (31.2%) | 11 (45.8%) | 13 (54.2%) | 0.066 |
| Diabetic nephropathy (%) | 47 (61.0%) | 29 (61.7%) | 18 (38.3%) | 0.881 |
| Age (year) | 56.5±11.3 | 55.6±12.2 | 58.0±9.5 | 0.799 |
| BMI | 25.7±4.2 | 25.9±4.3 | 25.5±4.2 | 0.706 |
| SBP (mmHg) | 150.3±23.8 | 149.5±23.0 | 151.5±25.3 | 0.657 |
| DBP (mmHg) | 84.2±11.2 | 84.6±10.4 | 83.4±12.4 | 0.775 |
| HbA1c (%) | 5.8±1.2 | 5.8±0.9 | 5.8±1.6 | 0.607 |
| PTH (pg/mL) | 259.4±192.8 | 253.7±176.0 | 268.4±219.3 | 0.418 |
| HB (g/L) | 89.1±18.1 | 89.7±18.5 | 88.2±17.5 | 0.726 |
| PLT(*10<sup>9</sup>/L) | 205.9±75.9 | 208.6±71.6 | 201.6±83.2 | 0.696 |
| TT (mmol/L) | 1.7±1.0 | 1.7±1.0 | 1.8±1.1 | 0.614 |
| ALB (g/L) | 33.2±9.0 | 33.4±10.5 | 32.8±6.2 | 0.496 |
| SCr (umol/L) | 671.8±246.4 | 649.6±239.6 | 706.5±256.9 | 0.326 |
| Ca (mmol/L) | 1.9±0.3 | 2.0±0.3 | 2.0±0.2 | 0.929 |
| P (mmol/L) | 1.7±0.4 | 1.6±0.4 | 1.8±0.4 | 0.004 |
| Ca&P product (mmol<sup>2</sup>/L<sup>2</sup>) | 3.3±1.0 | 3.1±1.0 | 3.6±1.1 | 0.078 |
| CRP (mg/L) | 10.1±16.4 | 10.1±17.2 | 10.2±15.4 | 0.807 |
| Diameter of cephalic vein (mm) | 2.6±0.6 | 2.6±0.5 | 2.5±0.7 | 0.764 |
| Diameter of radial artery (mm) | 2.5±0.4 | 2.6±0.3 | 2.5±0.5 | 0.392 |
| TC (mmol/L) | 4.4±1.3 | 4.1±1.0 | 4.9±1.7 | 0.024 |
| LDL(mmol/L) | 2.8±1.5 | 2.6±0.9 | 3.1±1.5 | 0.010 |

SBP: systolic blood pressure, DBP: diastolic blood pressure, HbA1c: glycosylated hemoglobin, TT:
total triglyceride, ALB: Albumin, SCr: creatinine, Ca: Calcium, P: Phosphorus, CRP: C-reactive
protein, TC: total cholesterol, LDL: Low density lipoprotein

## 3. AVF failure

Figures 1 to 4 shows the survival curves with respect to phosphorus, TC, LDL, and gender, respectively. Log rank tests suggested statistically significant differences in all survival curves, and the Log Rank P value was 0.007, 0.001, 0.012, 0.013, respectively. Therefore, phosphorus, TC, LDL, Gender, HbA1c and age were included in the multivariate cox proportional hazard regression, which revealed that hypercholesterolemia and hyperphosphatemia were independent risk factors for AVF dysfunction (P < 0.05) (Table 2).

**Table 2.** Multivariate Cox proportional hazard regression analysis.

| Risk factor | *P value* | RR value | 95%CI |  |
| --- | --- | --- | --- | --- |
|  |  |  | lower limit | upper limit |
| Phosphorus | 0.032 | 2.389 | 1.078 | 5.249 |
| TC | 0.043 | 3.115 | 1.037 | 9.360 |
| LDL | 0.589 | 1.288 | 0.514 | 3.229 |
| Gender | 0.278 | 1.573 | 0.694 | 3.567 |
| HbA1c | 0.641 | 0.796 | 0.305 | 2.076 |
| age | 0.399 | 1.460 | 0.606 | 3.517 |

**Fig. 1.**
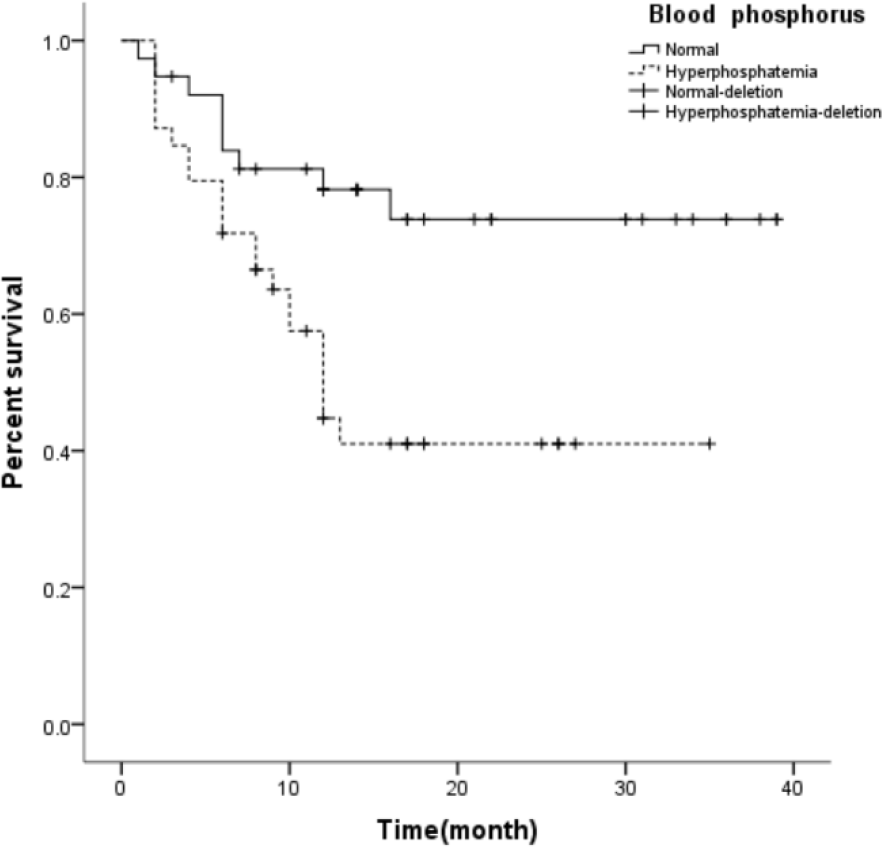
AVF survival curves for patients with hyperphosphatemia and normal blood phosphorus

**Fig. 2.**
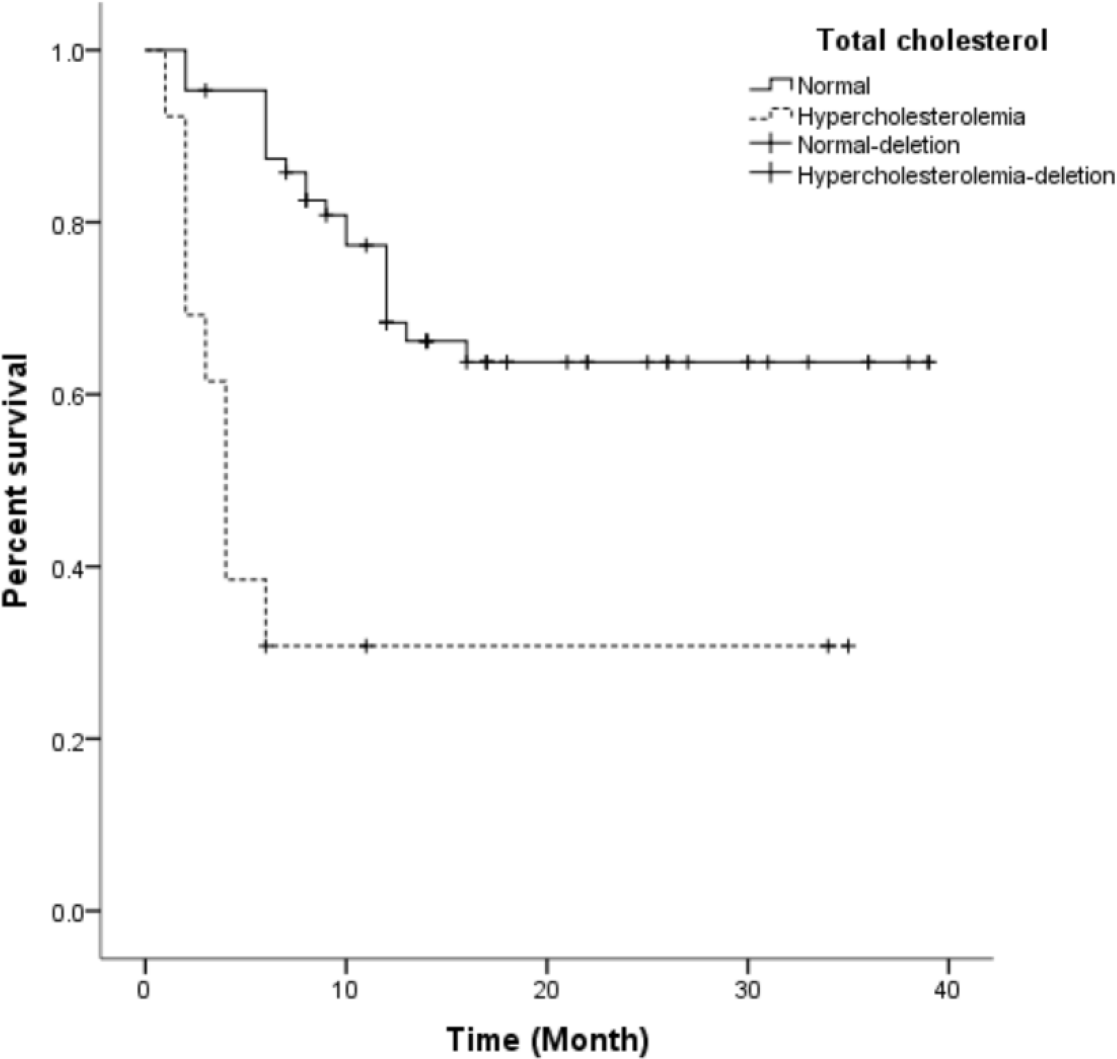
AVF survival curves for patients with hypercholesterolemia and normal TC

**Fig. 3.**
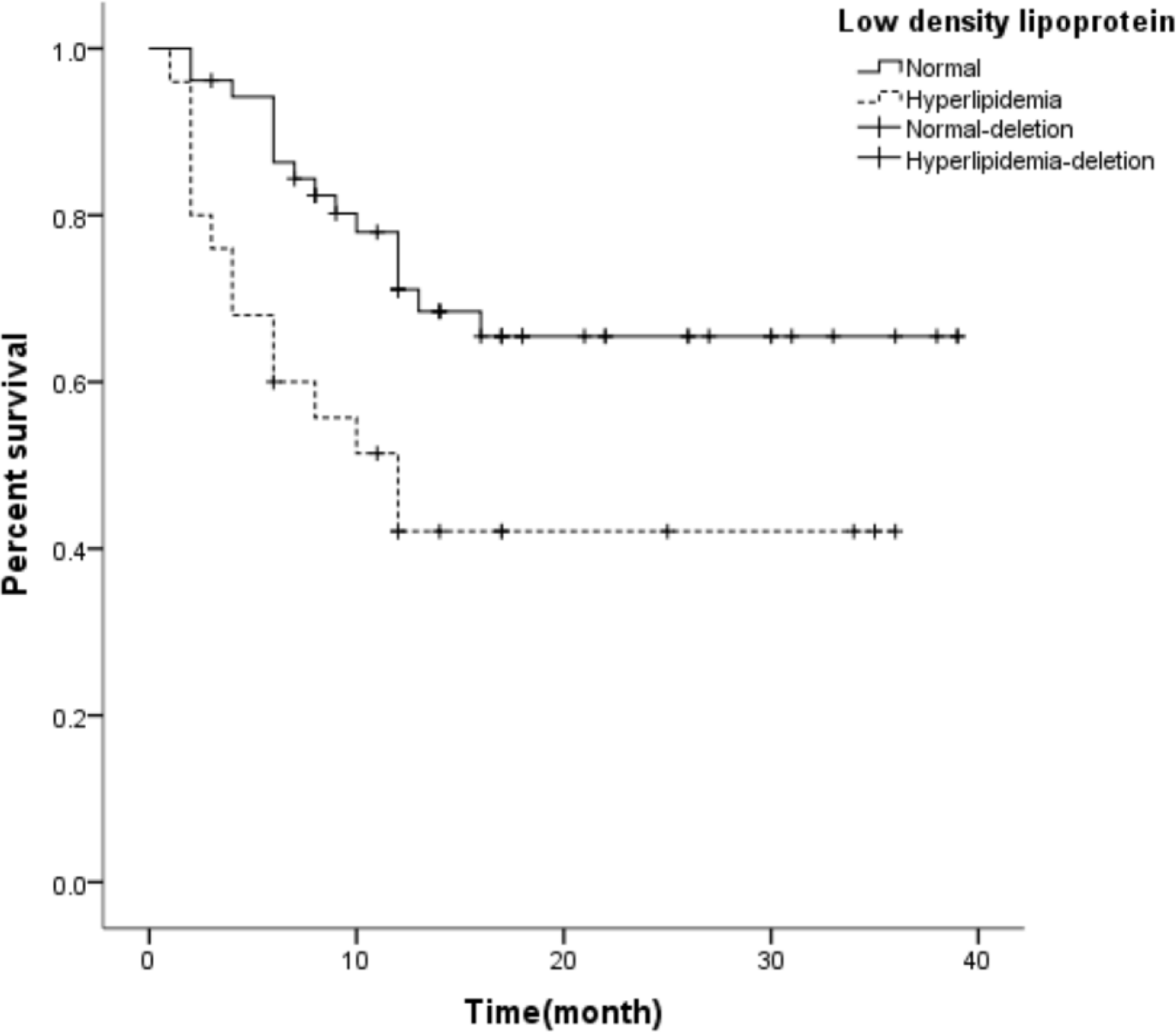
AVF survival curves for patients with increased LDL and normal LDL

**Fig. 4.**
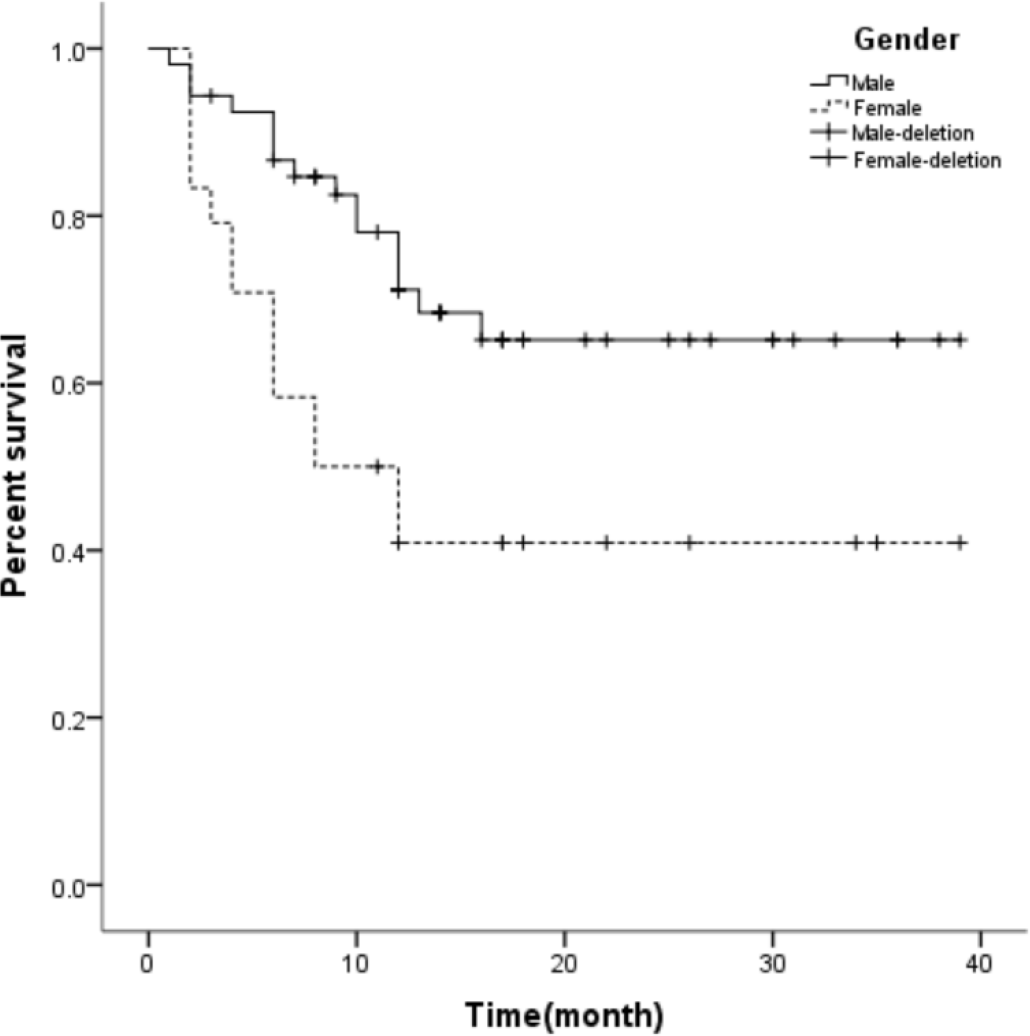
AVF survival curves for patients with male and female

## 4. Discussion

### Factors influencing development of AVF

Over the past 50 decades, there has been a growing consensus of “fistula first” principle in patients with MHD. The guidelines, published in 1997 (1) and revised in 2003 (2) and 2006 (3), together with the “fistula first” principle proposed in 2003 (4), suggests that AVF is the optimal choice. Although AVF is associated with thrombosis, blood steal syndrome and aneurysm, incidence of these complications are relatively low for AVF nowadays. Many factors, such as blood calcium, phosphorus and lipid (5), CRP (6), SCr; vascular diameter (7) and preoperative radial artery calcification (RAC) (8), can not only be caused by renal failure (9) but also affect the patency of AVFs. Although some studies have reported some of these risks, many have not been exhaustive.

Since the “fistula first” principle in 2003 (10), clinicians have increasingly preferred AVF for HMD. However, studies show that the AVF failure ranges between 15% and 22% (2,11). Currently, little emphasis has been putted on preoperative internal environment. Recent studies suggest that the internal environmental factors of AVFs are important (12). In this study, we found TC and phosphorus to be independent risk factors for AVF dysfunction.

### 4.1 Age

Most elderly patients present with other underlying diseases. As such, they may not harness optimal benefits from certain disease interventions, evidenced by short survival rate (13). In a separate but related study, it was found that the advantages of AVF over central venous catheter (CVC) gradual decreases with age. However, the disadvantages of AVF are gradually amplified. As such, CVC usage may be preferred in elderly patients. Meanwhile, the study shows that among the patients who have die of CKD, 35% of them were on AVF (14). Therefore, the huge loss is not worthy undisputedly embracing “fistula first”. Combined with findings of this study, age is not an independent risk factor for AVF dysfunction. Generally, age of patients does not affect the primary patency rate of AVF, but affect the long-term benefits. At the same time, from an economic point of view, AVF use for dialysis in elderly patients may be counterproductive. In a related study, K. hall et al. (15) established a Markov model from retrospective data to analyze the input-output ratio of patients with AVF, arteriovenous graft (AVG) and CVC. They found the input-output of AVF against AVG and CVC gradually declines with patients’ age. Thamer M et al. (16) also showed that in the whole age population, the average annual cost of maintaining AVF patency for 2.5 years is $7871 whereas that of patients with primary dysfunction is $13282 per year. So, the benefits of AVF in elderly patients are sub-optimal.

### 4.2 Serum total cholesterol (TC)

Blood lipid plays an important role in the patency of arteriovenous fistula. This research shows that TC is an independent risk factor for AVF dysfunction (17). Generally, high-density lipoprotein (HDL) is a protective factor for AVF dysfunction. Wang et al. (18) demonstrated that higher TT increases the risk of AVF dysfunction. In this research, TC was an independent risk factor for AVF, but TT was not. This discrepancy may however be attributed to our small sample size. Mechanistically, hyperlipidemia activates vascular endothelial growth factor A/matrix metalloproteinase-9 signaling pathway, which causes intimal hyperplasia and atherosclerosis (16).

### 4.3 Blood phosphorus

Many studies have implicated blood phosphorus for AVF dysfunction. Moon and Lee (19) showed that hyperphosphatemia (blood phosphorus > 5.5mg/ml) has a strong positive correlation with AVF dysfunction within 1 year of its induction. In a related study, Zhou Min and Lu Fangping (20) found that patients with hyperphosphatemia were more likely to develop AVF dysfunction than their normal counterparts. They found hyperphosphatemia to be an independent risk factor for AVF dysfunction, consistent with our findings. Continuous exploration of vascular access dysfunction has drawn interest in vascular calcification. Blood phosphorus has been implicated for vascular calcification. Here, prolonged hyperphosphatemia transforms vascular smooth muscle cells (VSMC) into osteoblast like cells (21). This induces calcium and phosphorus deposition on the inner wall of blood vessels, thus causing vascular calcification.

### 4.4 Serum creatinine

The current consensus is that AVF is the most preferred vascular access in dialysis patients. However, the timing of fistula establishment remains unclear. Y Wang, et al. (22) opined that before AVF surgery, vascular ultrasound and other examinations should be performed and evaluated, respectively. At the same time, Stolic et al. did not find SCr as an independent risk factor for AVF dysfunction (23). Based on our findings, SCr is not an independent risk factor for AVF dysfunction. In other words, the renal function (serum creatinine) at the initiation of dialysis does not strongly impact on the primary patency rate. As such, clinicians should not only consider the level of SCr, but also other factors before initiation of MHD.

### 4.5 Vessel diameter

This study also investigated the effect of vascular (arterial and venous) diameters on AVF primary dysfunction. Even though Misskey Jonathan and hamidzadeh Ramin (24) found radial arterial diameter (<2.1mm) to be an independent risk factors for AVF dysfunction. However, we report dissimilar findings: Vascular diameter not to be an risk factor for AVF dysfunction, probably due to strong emphasis on vascular diameter before surgery in our center.

## 5. Conclusion

Findings of this study implicate TC and phosphorus as independent risk factors for arteriovenous fistula dysfunction, and these factors can reduce the primary patency rate. Therefore, before the operation, patients should be assessed appropriately. Currently, there are many measures to correct hypercholesterolemia, among which the most commonly used is lipid-lowering drugs. And the results are satisfactory. However, it is relatively difficult to correct hyperphosphatemia. The main treatment measures were “3D principle”, including diet, dialysis and drugs (25). But the effect is unsatisfactory. In recent years, great progress has been made in the mechanism of hyperphosphatemia leading to intimal hyperplasia in AVFs. Blood phosphorus will induce phenotype transformation of VSMC, and eventually lead to intimal hyperplasia through complex signaling pathways (26). Therefore, to identify and block the signaling pathway may block intimal hyperplasia, which is very promising. The patients will be poised to benefit optimally after AVF.

## 6. Limitation

We acknowledge that our study analyzes data from a single center and may not be avoid of selection bias. Being a retrospective study with a small sample size, we cannot exclude other confounding. Due to the limitation of our dataset, 23 were lost to follow-up for various reasons, 10 received kidney transplant, and 11 died, which may lead to the deviation of the results. Power test was also performed, however, a larger sample size study is needed in the future. The relationship between a certain factor and AVF dysfunction needs further study as well.

## Data Availability

Anyone who wants to use the data in the text will need to be consented by the authors

## Funding

Beijing Hospitals Authority Youth Programme (QMS20200803 to C.Z)

## Footnote

### Reporting Checklist

The authors have completed the STROBE reporting checklist.

### Conflicts of Interest

All authors have completed the ICMJE uniform disclosure form. The authors have no conflicts of interest to declare.

### Ethical Statement

The authors are accountable for all aspects of the work in ensuring that questions related to the accuracy or integrity of any part of the work are appropriately investigated and resolved. The study protocol conformed to the ethical guidelines of the Declaration of Helsinki (as revised in 2013), and the Ethics Committee of Xuanwu Hospital approved this study (No.L YS[2020]018). Because of the retrospective nature of the research, the requirement for informed consent was waived.

## Notes

### Competing Interest Statement

The authors have declared no competing interest.

### Funding Statement

The author(s) received no financial support for the research, authorship, and/or publication of this article.

### Author Declarations

The study was approved by the ethics committee of Xuanwu Hospital, Capital Medical University

## Reference

[1] NKF-DOQI clinical practice guidelines for vascular access. National Kidney Foundation-Dialysis Outcomes Quality Initiative. Am J Kidney Dis 1997;30(4 Suppl 3):S150–S191.

[2] NKF-K/DOQI Clinical Practice Guidelines for Vascular Access: update 2000. Am J Kidney Dis 2001;37(1 Suppl 1):S137–S181.

[3] Vascular Access Work Group. Clinical practice guidelines for vascular access. Am J Kidney Dis 2006;48 Suppl 1:S248–S273.

[4] Hemodialysis Adequacy 2006 Work Group. Clinical practice guidelines for hemodialysis adequacy, update 2006. Am J Kidney Dis 2006;48 Suppl 1:S2–S90.

[5] C Ren, W Lu, X Yang, et al. Analysis on the application and the related factors of vascular access at initiation of hemodialysis in Shanghai. Chinese Journal of Blood Purification 2016, 15 (06): 344-347. China

[6] Zadeh M K, Mohammadipour S, Omrani Z. Correlation between CRP and early failure of arteriovenous fistula (AVF). Medical Journal of the Islamic Republic of Iran 2015, 29(1):219–219.

[7] Khavanin Zadeh M, Gholipour F, Naderpour Z, Porfakharan M. Relationship between vessel diameter and time to maturation of arteriovenous fistula for hemodialysis Access. Int J Nephrol 2012;2012:942950.

[8] Z Chen, H Zeng, F Huang. et al. Effect of radial artery calcification on survival of arteriovenous fistula and the patients in endstage renal disease patients. China Medical Abstracts(Internal Medicine) 2019,36(01):52. China

[9] Yan Y, Ye D, Yang L, et al. A meta-analysis of the association between diabetic patients and A VF failure in dialysis. Ren Fail 2018;40(1):379–383.

[10] National Kidney Foundation (2015). KDOQI Clinical Practice Guideline for Hemodialysis Adequacy: 2015 update. American journal of kidney diseases : the official journal of the National Kidney Foundation,66(5), 884–930.

[11] He C, Zhang X. Renal replacement therapy. Shanghai Science and Technology Education Press, 2005.152–154.

[12] Wen M, Li Z, Li J, et al. Risk factors for primary arteriovenous fistula dysfunction in hemodialysis patients: A retrospective survival analysis in multiple medical centers. Blood Purification 2019:1–7.

[13] Lee Timmy, Allon Michael. Reassessing Recommendations for Choice of Vascular Access. Clin J Am Soc Nephrol 2017, 12: 865–867.

[14] Woo K, Lok CE. New Insights into dialysis vascular access: What is the optimal vascular access type and timing of access creation in CKD and dialysis patients? Clin J Am Soc Nephrol 2016;11(8):1487–1494.

[15] Hall Rasheeda K, Myers Evan R, Rosas Sylvia E et al. Choice of hemodialysis access in older adults: A cost-effectiveness analysis. Clin J Am Soc Nephrol 2017;12(6):947–954.

[16] Thamer M, Lee TC, Wasse H, et al. Medicare Costs Associated With Arteriovenous Fistulas Among US Hemodialysis Patients. Am J Kidney Dis 2018;72(1):10–18.

[17] T Cui, R Zhang, F Liu, et al. Effect of diabetes mellitus on early dysfunction of arteriovenous fistula in patients with end-stage renal disease. Journal of Sichuan University (Medical Edition) 2012,43 (03): 438–441. China

[18] Z Li, W Wang, Y Jiang, et al. Analysis of influencing factors of failure of autogenous arteriovenous fistula in maintenance hemodialysis patients. Chinese Journal of Integrated Traditional and Western Nephrology. 2014,15(11):961-964. 292 China

[19] Moon JY, Lee HM, Lee SH, et al. Hyperphosphatemia is associated with patency loss of arteriovenous fistula after 1 year of hemodialysis. Kidney Res Clin Pract 2015;34(1):41–46.

[20] M Zhou, F Lu. Effect of hyperphosphatemia on patency rate after reoperation of autogenous arteriovenous fistula dysfunction. Chinese Journal of medicine 2018,98 (42): 3406–3410. China

[21] Shimokado A, Sun Y, Nakanishi M, et al. Smad3 plays an inhibitory role in phosphate-induced vascular smooth muscle cell calcification. Exp Mol Pathol 2014;97(3):458–464.

[22] Y Wang, C Ye, Z Jin. Chinese expert consensus on vascular access for hemodialysis (1st Edition). Chinese Journal of Blood Purification 2014, 13 (8), 549–558. China

[23] Stolic Radojica V, Trajkovic Goran Z,Kostic Mirjana M et al. Factors affecting the patency of arteriovenous fistulas for hemodialysis: Single center 307 experience.[J]. Hemodial Int, 2018, 22: 328–334.

[24] Misskey J, Hamidizadeh R, Faulds J, et al. Influence of artery and vein diameters on autogenous arteriovenous access patency. J Vasc Surg 2020;71(1):158–172.

[25] Wei Chen, Xueqing Yu. Challenges and advances in the management of hyperphosphatemia in dialysis patients with chronic kidney disease. [J]. Chinese Journal of Nephrology, 2018, 34(11): 867–871.

[26] Hénaut L, Mary A, Chillon JM, et al. The Impact of Uremic Toxins on Vascular Smooth Muscle Cell Function. Toxins (Basel). 2018;10(6):218.

